# Contribution of post-TB sequelae to life-years and quality-adjusted life-years lost due to TB disease in the United States, 2015-2019

**DOI:** 10.1101/2024.10.25.24316143

**Authors:** Nicolas A. Menzies, Suzanne M. Marks, Yuli L. Hsieh, Nicole A. Swartwood, Garrett R. Beeler Asay, Jacek Skarbinski, C. Robert Horsburgh, Ted Cohen

**Affiliations:** Department of Global Health and Population, Harvard T.H. Chan School of Public Health, Boston MA, USA; Center for Health Decision Science, Harvard T.H. Chan School of Public Health, Boston, MA, USA; CDC Division of Tuberculosis Elimination, U.S. Centers for Disease Control and Prevention, Atlanta, GA, USA; Interfaculty Initiatives in Health Policy, Harvard University, Cambridge, USA; Department of Infectious Diseases, Oakland Medical Center, Kaiser Permanente Northern California, Oakland, CA, USA; Departments of Epidemiology, Biostatistics, Global Health and Medicine, Boston University Schools of Public Health and Medicine, Boston, MA, USA; Department of Epidemiology of Microbial Diseases, Yale School of Public Health, New Haven, CT, USA

**Author notes:** **Corresponding Author:** Nicolas A. Menzies, Department of Global Health and Population, Harvard T.H. Chan School of Public Health, 665 Huntington Ave, Boston, MA 02115, USA.

## Abstract

**Background:** Individuals surviving TB disease may experience chronic sequelae that reduce survival and quality-of-life. These post-TB sequalae are not generally considered in estimates of the health impact of TB disease. We estimated reductions in life expectancy and quality-adjusted life expectancy for individuals developing TB disease in the United States, including post-TB sequelae.

**Methods:** We extracted national surveillance data on individuals diagnosed with TB during 2015-2019, including demographics, vital status at diagnosis, treatment duration, treatment outcome, and co-prevalent conditions. Using a mathematical model we simulated life expectancy and quality-adjusted life-years (QALYs) for the TB cohort, as compared to a no-TB counterfactual. We disaggregated results to report the proportion due to post-TB sequelae, and stratified outcomes by age, sex, and race.

**Findings:** Estimated life expectancy after TB diagnosis was 30.3 (95% uncertainty interval: 29.9, 30.7) years for the TB cohort versus 32.3 (31.9, 32.7) without TB, a difference of 2.03 (1.84, 2.21) years and 1.93 (1.69, 2.18) QALYs. Life-years lost were greatest for 65-74-year-olds versus other age groups, for men versus women, and for American Indian or Alaska Native individuals versus persons from other race/ethnicities. Overall, 41% (35, 46) of life-years and 48% (42, 54) of QALYs lost were estimated to result from post-TB sequelae.

**Interpretation:** In the United States, a substantial fraction of the life-years and QALYs lost from TB are attributable to post-TB sequelae. Evidence is needed on approaches to prevent and repair post-TB lung damage, in the context of frequent co-prevalent health conditions.

**Funding:** CDC.

**Research in Context:** *Evidence before this study:* Individuals surviving TB disease may experience reduced quality-of-life and elevated mortality rates, due to sequelae of the TB episode and pre-existing factors. We reviewed published literature to identify studies quantifying the impact of post-TB sequelae on life expectancy or other summary measures of health attainment. Using the following search terms: (tuberculosis) AND (“post-TB” OR “post-tuberculosis” OR “sequelae” OR “TB survivor” OR “pulmonary impairment” OR “delayed mortality”) AND (“life expectancy” OR “QALYs” OR “life years” OR “DALYs” OR “years of life lost”), we searched PubMed since inception until October 8 2024, without language restriction. Of the studies identified, most estimated health losses attributable to TB and post-TB in high-burden settings. Studies conducted in the United States reported results for specific geographic areas or trial populations, with estimates of the average life-years lost per TB case ranging from 1.5 to 7.0 years.

*Added value of this study:* For individuals developing TB in the United States, average life expectancy after TB diagnosis was estimated to be 30.3 (95% uncertainty interval: 29.9, 30.7) years, as compared to 32.3 (31.9, 32.7) years under a counterfactual scenario that estimated lifetime outcomes without TB. On average, TB was estimated to reduce life expectancy by 2.03 (1.84, 2.21) years, or 1.93 (1.69, 2.18) quality-adjusted life years (QALYs). Overall, 41% (35, 46) of life years lost and 48% (42, 54) of QALYs lost were from post-TB sequelae. Per person developing TB, TB-attributable reductions in life expectancy were greatest for 65-74-year-olds versus other age groups, for men versus women, and for American Indian/Alaska Native individuals versus other race/ethnicities.

*Implications of all the available evidence:* In this high-income setting with substantial healthcare resources, TB still represents a major health risk for those who develop the disease. Even for individuals who successfully complete TB treatment, lifetime health outcomes are poorer than for people who never had TB, and almost half of the QALYs lost from TB result from post-TB sequelae.

## Introduction

Globally, an estimated 10.6 million individuals developed tuberculosis (TB) in 2022, and 1.3 million died with the disease.^1^ Even after completing treatment, many TB survivors experience extended post-TB morbidity, and observational studies have documented substantially higher mortality rates among TB survivors compared to matched controls from similar demographic groups.^2,3^ Accumulating evidence from high-burden settings suggests that TB survivors are at risk of lifelong disability, reduced economic productivity, and a range of physical, psychological and social sequalae.^4–7^ For low TB incidence settings like the United States there is more limited evidence on the long-term implications of post-TB. However, evidence from several empirical studies suggests that for TB survivors in these settings the health consequences may be comparable to the effects reported in high TB incidence settings.^8–11^

Estimates of the health consequences of post-TB lung disease are needed for a comprehensive assessment of the impact of TB on lifetime health outcomes. Published global estimates that have estimated the population health impact of TB have generally restricted the focus of analysis to morbidity and mortality occurring during the TB disease episode, and excluded post-TB sequelae.^12^ Understanding the lifetime consequences of TB is difficult, as the causal effect of TB must be separated from the impact of pre-existing health conditions, behaviors, and living conditions (healthcare access, economics, education, community context, and built environment) that differ between individuals who develop TB and the general population.

Studies that have attempted to quantify the health losses experienced by TB survivors have primarily been conducted at the global level, or focused on high TB incidence settings.^13–16^

In this study we estimated the population health impact—quantified as life-years and quality-adjusted life-years lost—due to TB disease occurring in the United States, inclusive of post-TB sequelae among TB survivors. To do so, we analyzed nationally representative data on U.S. individuals diagnosed with TB disease during 2015-2019, and simulated survival and quality-of-life for each individual compared to a counterfactual scenario assuming the individual had not developed TB. We report results for the cohort overall as well as stratified by key demographic factors. We also disaggregate overall TB-attributable reductions in life expectancy and quality-adjusted life expectancy to estimate the proportions resulting from TB and post-TB sequalae respectively.

## Methods

### Study cohort

The National TB Surveillance System (NTSS) includes data on all U.S. individuals diagnosed with TB disease.^17^ We extracted NTSS data (including outcomes reported to CDC as of July 8, 2023) for individuals diagnosed with TB during 2015–2019 and retained variables describing notification year, sex, age at diagnosis, self-described race/ethnicity, vital status at diagnosis, treatment duration, dates of TB treatment initiation and/or death, and treatment outcome. We also extracted variables recording the presence of co-morbidities, behaviors, and social factors (hereafter ‘co-prevalent conditions’) associated with elevated mortality rates or lower quality-of-life (e.g., HIV, diabetes, end stage renal disease or chronic renal failure [ESRD], solid organ transplant receipt, receipt of immunosuppressive medications, multidrug-resistant [MDR]-TB, excess alcohol use, injection drug use, and experiencing homelessness at the time of TB diagnosis). We used multiple imputation to impute missing values (see Supplementary Methods).^18^

### Survival of the TB episode

We created a binary variable representing whether or not an individual survived the TB episode. Individuals who completed treatment were assumed to have survived the TB episode. Individuals recorded as dead at diagnosis, or having died during treatment, were coded as having not survived the TB episode. Among individuals not meeting one of these criteria, individuals with a recorded death date and no treatment initiation date were assumed to be dead at diagnosis. Similarly, individuals with a recorded death date occurring after a recorded treatment initiation date were assumed to have died during treatment. Survival of the TB episode was unknown for individuals who did not meet one of these criteria (i.e., individuals who were alive at diagnosis but either refused treatment or discontinued treatment before completion). We treated these instances as missing values and imputed them using the multiple imputation approach used for other missing values. If this resulted in a death being imputed for a given individual, this death was assumed to occur at the recorded time of treatment discontinuation.

### Simulation model

We constructed a mathematical model following individuals with incident TB disease during their disease episode and over their remaining lifetimes. The model was operationalized with a monthly timestep, considering the symptomatic pre-treatment period (conceptualized as the period before TB diagnosis with elevated TB mortality and reduced quality-of-life due to TB), the treatment period, and the post-treatment period.

*Pre-treatment period*: Each individual in the study cohort was represented in the model, parameterized with the characteristics recorded in the NTSS data. The model was initialized at the beginning of the symptomatic pre-treatment period, which was assumed to begin 2 months before TB diagnosis.^19^ Deaths during the pre-treatment period were based on the reported values for dead at diagnosis in NTSS. These pre-treatment deaths were assumed to be distributed uniformly over the pre-treatment period. We used utility weights to quantify the quality-of-life associated with TB and co-prevalent conditions (Table 1). Individuals were also assumed to experience age-specific background utility reductions.^20^ We used a multiplicative approach to combine utility weights from multiple conditions (equation in Supplement).

**Table 1:** Parameter values and sources. * Mortality rate ratios and utility weights for individuals receiving immunosuppressive medications (TNF-alpha inhibitors, systemic corticosteroids) based on rheumatoid arthritis. ^†^ Value based on standard gamble utility estimates for individuals receiving TB treatment compared to control, at treatment initiation. ^‡^ Value based on standard gamble utility estimates for individuals receiving TB treatment compared to control, averaged over months 2-6 after diagnosis.

| Parameter definition | Parameter value<br>(mean, 95% interval) | Source |
| --- | --- | --- |
| Background mortality rates | Stratified by age, sex, and race/ethnicity, no uncertainty | 22 |
| Mortality for individuals with TB during pre-treatment and treatment period | As reported in NTSS. Uncertainty in missing values represented by multiple imputation. | 17 |
| Mortality rate ratio for post TB among TB survivors compared with people with no TB | 1.78 (1.61, 1.98) | 8 |
| Mortality rate ratios for co-prevalent conditions: |  |  |
| *Excess alcohol use* | 1.47 (1.28, 1.68) | 39 |
| *HIV infection* | 2.71 (2.17, 3.25) | 40 |
| *Experiencing homelessness* | 2.80 (2.10, 3.50) | 41 |
| *Injection drug use* | 2.40 (1.65, 3.48) | 42 |
| *Organ transplant recipient* | 4.86 (3.65, 6.08) | 43 |
| *Diabetes* | 1.80 (1.35, 2.25) | 44 |
| *End-stage renal disease* | 12.0 (9.0, 15.0) | 45 |
| *Immunosuppressive medication\** | 1.54 (1.41, 1.67) | 46 |
| Background utility weight | Stratified by age | 20 |
| Utility weight during pre-treatment period and first month of treatment<sup>†</sup> | 0.750 (0.660, 0.830) | 21 |
| Utility weight during treatment period, after first month<sup>‡</sup> | 0.890 (0.780, 0.980) | 21 |
| Utility weight during post-treatment period | 0.988 (0.982, 0.994) | 13 |
| Utility weight for co-prevalent conditions: |  |  |
| *HIV infection* | 0.950 (0.900, 0.980) | 47,48 |
| *Organ transplant recipient* | 0.820 (0.740, 0.900) | 49 |
| *Diabetes* | 0.963 (0.950, 0.976) | 50 |
| *End-stage renal disease* | 0.750 (0.650, 0.850) | 51,52 |
| *Immunosuppressive medication\** | 0.910 (0.902, 0.918) | 53 |

*Treatment period*: The length of treatment was based on the recorded treatment duration, rounded to the nearest month. Deaths during treatment were assumed to occur at the recorded date of death if available, or otherwise at the end of the recorded treatment duration. We assumed that deaths recorded in NTSS would include those due to non-TB causes and did not assume additional mortality from other causes during pre-treatment and treatment periods. Quality-of-life during treatment was based on published utility values (Table 1).^21^

*Post-treatment period*: Mortality during the post-treatment period is not recorded in NTSS, and instead we modelled mortality over this period as a function of background mortality, elevated mortality due to co-prevalent conditions, and elevated post-TB mortality. Background mortality rates were taken from national sex-and race/ethnicity-stratified lifetables for 2019.^22^ Mortality rate ratios associated with co-prevalent conditions were extracted from the published literature (Table 1). For individuals with multiple co-prevalent conditions we assumed their effect on overall mortality would combine additively (equation in Supplement).^23,24^ The mortality rate ratio for post-TB (1.78 (1.61, 1.98)) was derived from a retrospective cohort study of TB survivors compared to controls with no TB diagnosis, matched on age, sex, race, year of diagnosis, and a range of pre-existing conditions.^8^ We assumed the post-TB mortality rate ratio applied over the average follow-up in the source study (6.8 years) and subsequently returned to 1.0. For the post-treatment period we applied a utility weight of 0.988 (0.982, 0.994), based on country-specific results from a global study of the burden of disease attributable to post-TB.^13^

### Counterfactual scenarios

We compared results for the TB cohort to results for a ‘non-TB’ counterfactual scenario, in which we simulated future survival and quality-of-life for a cohort of individuals similar to the study cohort in all respects (same distribution across age group, sex, race/ethnicity, and co-prevalent conditions) but without TB. For these individuals, mortality rates were based on background mortality rates plus the combined mortality rate ratio associated with other conditions (Table 1). Utility weights were based on age-specific background utility and utility weights for co-prevalent conditions. The difference in outcomes between TB and no-TB scenarios was used to quantify the lifetime consequences of TB.

We also estimated outcomes for a third scenario (‘TB-episode-only’), which was based on the TB scenario but omitting the post-TB mortality rate ratio and utility reduction over the post-treatment period. We used the results of this scenario to disaggregate overall health losses associated with TB into the proportion occurring during the TB episode (combining pre-treatment and treatment periods) and the proportion associated with post-TB (post-treatment period).

### Outcomes

Health outcomes included life expectancy and quality-adjusted life expectancy (QALE). We also calculated the reduction in life expectancy and QALE due to TB, computed as the incremental difference in outcomes between the no-TB scenario and the TB scenario. Health losses occurring during the TB episode were calculated as the incremental difference between no-TB and TB-episode-only scenarios. Post-TB health losses were calculated as the incremental difference between TB-episode-only and TB scenarios. We report average and total reductions in life expectancy and quality-adjusted life expectancy in the study cohort, and also divided total life expectancy losses by the number of person-years at risk of TB to report life-years lost per million person-years. We calculated 95% uncertainty intervals using Monte Carlo simulation (see Supplement).

We report results for the overall study cohort as well as stratified by covariates: notification year (2015-2019), sex (male, female), age group (0-4, 5-14, 15-24, 25-34, 35-44, 45-54, 55-64, 65-74, 75-84, and 85+ years) and race/ethnicity (American Indian or Alaska Native (AIAN), Hispanic, Non-Hispanic Asian (Asian), Non-Hispanic Black (Black), Non-Hispanic White (White), and Native Hawaiian or Other Pacific Islander (NHPI)).

We fit linear regression models to the individual-level estimates of life-years lost and QALYs lost due to TB, to report the differences in these outcomes as a function of notification year, sex, age group and race/ethnicity, controlling for each of these demographic factors simultaneously. Regression models were fit for each parameter set, and the results combined using Rubin’s rule.^25^

### Sensitivity analysis

We conducted sensitivity analyses to test the robustness of results to imputation assumptions: first, we re-estimated results from the subset of records with no missing data; and second, we re-estimated results using the most optimistic and most pessimistic assumptions for missing data on TB survival (equivalent to assuming all individuals with missing survival data survived, and died, respectively).

## Results

There were 45,738 NTSS-reported TB cases in the United States during 2015-2019 (Table 2). Of this total, 61% were male and the average age was 48 (interquartile range: 31-65). The most common race/ethnicities were Asian (35%), Hispanic (29%), and Black (21%), and 40% had at least one co-prevalent condition (Table S1). Overall, 5.9% (n=2710) of individuals were missing data on TB survival, 13.8% (n=6323) were missing data on other covariates used in the analysis, and 18.0% (n= 8241) had any missing data (Figure S1 shows the number of missing values for each variable). After imputation of missing data, 2.4% of individuals were classified as dead at diagnosis, 7.0% died during treatment, and 90.6% survived the disease episode.

**Table 2.** Study cohort and outcomes of TB disease episode, by notification year and demographic stratum. Results include imputed values for missing demographic variables and treatment outcomes (Figure S1 shows missingness for each variable). Values in parentheses represent the range of values across 100 multiply-imputed datasets. ‘AIAN’ = American Indian or Alaska Native. ‘Asian’ = Non-Hispanic Asian. ‘Black’ = Non-Hispanic Black. ‘White’ = Non-Hispanic White. ‘NHPI’ = Native Hawaiian or Other Pacific Islander. ‘Other’ = Non-Hispanic other race or people who report more than one race.

|  | Total cohort | Outcome of TB episode |  |  |
| --- | --- | --- | --- | --- |
|  |  | Died before diagnosis | Died during treatment | Survived |
| Overall | 45,738 (45,738, 45,738) | 1,086 (1,070, 1,105) | 3,201 (3,163, 3,235) | 41,451 (41,409, 41,494) |
| Notification year |  |  |  |  |
| 2015 | 9,538 (9,538, 9,538) | 222 (215, 230) | 610 (597, 627) | 8,706 (8,692, 8,722) |
| 2016 | 9,239 (9,239, 9,239) | 227 (218, 234) | 643 (630, 658) | 8,369 (8,350, 8,391) |
| 2017 | 9,069 (9,069, 9,069) | 181 (175, 190) | 644 (632, 659) | 8,244 (8,230, 8,257) |
| 2018 | 8,997 (8,997, 8,997) | 216 (207, 230) | 617 (604, 629) | 8,165 (8,152, 8,182) |
| 2019 | 8,895 (8,895, 8,895) | 240 (232, 250) | 688 (676, 704) | 7,968 (7,952, 7,981) |
| Sex |  |  |  |  |
| Men | 27,754 (27,752, 27,757) | 694 (672, 705) | 2,139 (2,116, 2,161) | 24,921 (24,896, 24,958) |
| Women | 17,984 (17,981, 17,986) | 392 (382, 406) | 1,062 (1,042, 1,081) | 16,530 (16,507, 16,554) |
| Age group |  |  |  |  |
| 0-4 years | 1,094 (1,094, 1,096) | 6 (6, 7) | 8 (8, 10) | 1,080 (1,078, 1,082) |
| 5-14 years | 900 (900, 901) | 0 (0, 1) | 1 (1, 2) | 899 (898, 900) |
| 15-24 years | 4,431 (4,431, 4,433) | 12 (11, 14) | 36 (33, 40) | 4,384 (4,378, 4,388) |
| 25-34 years | 7,444 (7,443, 7,446) | 37 (34, 45) | 85 (80, 91) | 7,322 (7,309, 7,328) |
| 35-44 years | 6,329 (6,329, 6,331) | 58 (54, 65) | 153 (147, 160) | 6,118 (6,109, 6,125) |
| 45-54 years | 6,618 (6,618, 6,620) | 125 (119, 134) | 314 (305, 324) | 6,180 (6,170, 6,192) |
| 55-64 years | 7,339 (7,339, 7,342) | 194 (188, 202) | 595 (581, 615) | 6,550 (6,531, 6,566) |
| 65-74 years | 5,687 (5,687, 5,689) | 222 (212, 231) | 703 (689, 719) | 4,762 (4,747, 4,779) |
| 75-84 years | 4,149 (4,149, 4,151) | 266 (256, 281) | 778 (765, 792) | 3,105 (3,089, 3,125) |
| 85+ years | 1,745 (1,745, 1,747) | 166 (161, 173) | 528 (515, 539) | 1,051 (1,038, 1,064) |
| Race/ethnicity |  |  |  |  |
| AIAN | 533 (532, 536) | 20 (19, 24) | 42 (41, 46) | 471 (466, 474) |
| Hispanic | 13,190 (13,176, 13,203) | 278 (269, 289) | 762 (746, 780) | 12,150 (12,122, 12,172) |
| Asian | 16,188 (16,176, 16,203) | 313 (303, 322) | 1,180 (1,166, 1,196) | 14,695 (14,679, 14,716) |
| Black | 9,467 (9,455, 9,478) | 247 (237, 257) | 625 (610, 644) | 8,596 (8,572, 8,616) |
| White | 5,567 (5,561, 5,573) | 221 (214, 229) | 551 (540, 567) | 4,796 (4,773, 4,809) |
| NHPI | 506 (504, 510) | 4 (4, 5) | 23 (22, 25) | 479 (476, 483) |
| Other | 287 (286, 290) | 3 (3, 5) | 18 (17, 21) | 265 (262, 269) |

### Estimated outcomes for TB and no-TB cohorts

We estimated mean life expectancy for individuals in the TB cohort to be 30.3 (95% uncertainty interval: 29.9, 30.7) years from the development of symptomatic TB, and 32.3 (31.9, 32.7) for matched individuals without TB, with TB estimated to have reduced average life expectancy by 6.3% (5.7, 6.8). The relative reduction in life expectancy varied across demographic strata and was greatest for older individuals (Table S2). Quality-adjusted life expectancy was estimated to be 24.5 (23.1, 25.7) years for the TB cohort and 26.4 (24.9, 27.6) years for the no-TB counterfactual, equivalent to a 7.3% (6.5, 8.1) reduction attributable to TB (Table S3).

Table 3 reports the reductions in average life expectancy due to TB. Across the cohort, TB was estimated to reduce life expectancy by 2.03 (1.84, 2.21) years. Estimated reductions in life expectancy were greatest for 65-74-year-olds (3.04 (2.76, 3.32) years) and smallest for 5-14-year-olds (0.21 (0.18, 0.28) years), and larger for men compared to women (2.15 (1.96, 2.35) vs. 1.83 (1.67, 1.99) years). The AIAN population had the largest estimated life expectancy loss (2.83 (2.54, 3.13)) compared to other race/ethnicity populations. Across the cohort, TB was estimated to reduce quality-adjusted life expectancy by 1.93 (1.69, 2.18) QALYs, with similar patterns across demographic strata as for life expectancy.

**Table 3.** Average reduction in life expectancy and quality-adjusted life expectancy for individuals developing TB, by demographic group. Values in parentheses represent 95% uncertainty intervals. QALE = quality-adjusted life expectancy. ‘AIAN’ = American Indian or Alaska Native. ‘Asian’ = Non-Hispanic Asian. ‘Black’ = Non-Hispanic Black. ‘White’ = Non-Hispanic White. ‘NHPI’ = Native Hawaiian or Other Pacific Islander. ‘Other’ = Non-Hispanic other race or people who report more than one race.

|  | Reduction in life expectancy |  |  | Reduction in quality-adjusted life expectancy |  |  |
| --- | --- | --- | --- | --- | --- | --- |
|  | TB episode | Post-TB | Total | TB episode | Post-TB | Total |
| Overall | 1.19 (1.15, 1.24) | 0.83 (0.65, 1.02) | 2.03 (1.84, 2.21) | 1.00 (0.92, 1.10) | 0.93 (0.72, 1.15) | 1.93 (1.69, 2.18) |
| Sex |  |  |  |  |  |  |
| Men | 1.24 (1.20, 1.29) | 0.91 (0.72, 1.11) | 2.15 (1.96, 2.35) | 1.04 (0.95, 1.14) | 0.96 (0.76, 1.19) | 2.00 (1.76, 2.26) |
| Women | 1.12 (1.08, 1.16) | 0.71 (0.56, 0.87) | 1.83 (1.67, 1.99) | 0.94 (0.86, 1.04) | 0.87 (0.66, 1.09) | 1.81 (1.58, 2.06) |
| Age group |  |  |  |  |  |  |
| 0-4 years | 0.92 (0.89, 0.98) | 0.07 (0.05, 0.08) | 0.99 (0.95, 1.05) | 0.90 (0.83, 1.00) | 0.85 (0.50, 1.29) | 1.75 (1.39, 2.21) |
| 5-14 years | 0.08 (0.07, 0.15) | 0.13 (0.10, 0.17) | 0.21 (0.18, 0.28) | 0.16 (0.11, 0.25) | 0.81 (0.51, 1.21) | 0.98 (0.66, 1.38) |
| 15-24 years | 0.54 (0.50, 0.60) | 0.35 (0.27, 0.44) | 0.89 (0.80, 0.99) | 0.55 (0.48, 0.64) | 0.87 (0.61, 1.21) | 1.42 (1.13, 1.76) |
| 25-34 years | 0.61 (0.57, 0.65) | 0.43 (0.33, 0.53) | 1.04 (0.94, 1.15) | 0.60 (0.53, 0.69) | 0.83 (0.59, 1.10) | 1.42 (1.17, 1.71) |
| 35-44 years | 0.99 (0.95, 1.04) | 0.60 (0.46, 0.75) | 1.59 (1.45, 1.74) | 0.89 (0.82, 0.98) | 0.83 (0.62, 1.06) | 1.72 (1.50, 1.97) |
| 45-54 years | 1.39 (1.33, 1.46) | 0.95 (0.74, 1.18) | 2.34 (2.12, 2.57) | 1.17 (1.08, 1.28) | 0.98 (0.77, 1.20) | 2.15 (1.90, 2.42) |
| 55-64 years | 1.60 (1.53, 1.68) | 1.22 (0.96, 1.51) | 2.83 (2.56, 3.10) | 1.30 (1.18, 1.41) | 1.09 (0.86, 1.32) | 2.38 (2.11, 2.68) |
| 65-74 years | 1.75 (1.67, 1.84) | 1.29 (1.01, 1.57) | 3.04 (2.76, 3.32) | 1.37 (1.24, 1.50) | 1.04 (0.83, 1.27) | 2.41 (2.11, 2.70) |
| 75-84 years | 1.68 (1.60, 1.77) | 1.27 (1.02, 1.52) | 2.95 (2.67, 3.23) | 1.29 (1.16, 1.42) | 0.96 (0.77, 1.16) | 2.25 (1.96, 2.52) |
| 85+ years | 1.31 (1.25, 1.39) | 0.87 (0.72, 1.01) | 2.19 (2.00, 2.36) | 0.99 (0.88, 1.10) | 0.63 (0.50, 0.75) | 1.61 (1.41, 1.82) |
| Race/ethnicity |  |  |  |  |  |  |
| AIAN | 1.53 (1.42, 1.66) | 1.30 (1.03, 1.58) | 2.83 (2.54, 3.13) | 1.29 (1.18, 1.42) | 1.27 (1.02, 1.54) | 2.56 (2.26, 2.87) |
| Hispanic | 1.22 (1.17, 1.27) | 0.77 (0.60, 0.95) | 1.99 (1.81, 2.17) | 1.04 (0.95, 1.13) | 0.92 (0.70, 1.14) | 1.95 (1.71, 2.21) |
| Asian | 1.14 (1.10, 1.19) | 0.66 (0.52, 0.81) | 1.80 (1.66, 1.95) | 0.94 (0.86, 1.04) | 0.79 (0.60, 0.99) | 1.73 (1.51, 1.97) |
| Black | 1.12 (1.07, 1.18) | 1.07 (0.84, 1.30) | 2.19 (1.95, 2.44) | 0.97 (0.89, 1.07) | 1.11 (0.88, 1.36) | 2.08 (1.82, 2.36) |
| White | 1.37 (1.32, 1.42) | 1.06 (0.84, 1.29) | 2.43 (2.20, 2.66) | 1.12 (1.03, 1.22) | 1.00 (0.80, 1.22) | 2.13 (1.87, 2.38) |
| NHPI | 1.04 (0.96, 1.19) | 0.66 (0.51, 0.81) | 1.70 (1.54, 1.88) | 0.91 (0.81, 1.04) | 0.92 (0.69, 1.18) | 1.83 (1.58, 2.12) |
| Other | 1.15 (1.04, 1.43) | 0.74 (0.57, 0.90) | 1.89 (1.69, 2.19) | 1.02 (0.89, 1.25) | 0.92 (0.70, 1.15) | 1.93 (1.67, 2.24) |

Of the total reduction in life expectancy, an estimated 1.19 (1.15, 1.24) years (59% (54, 65) of total life-years lost) were lost due to deaths during the TB episode, and 0.83 (0.65, 1.02) years (41% (35, 46)) were lost due to elevated post-TB mortality. For quality-adjusted life expectancy individuals with TB were estimated to lose 1.00 (0.92, 1.10) QALYs during the TB episode (52% (46, 58) of total QALYs lost) and 0.93 (0.72, 1.15) QALYs during post-TB (48% (42, 54)).

We estimated 93,000 (84,000, 101,000) life-years lost in the study cohort due to TB, equivalent to 57 (52, 62) life-years lost per million person-years at risk (Table S4). Figure 1 shows how the number of life-years lost was distributed by age and race/ethnicity. While the absolute number of life-years lost was greatest for 55-64-year-olds, the greatest losses per million person-years were in 75-84-year-olds. The greatest number of life-years lost per million person-years was estimated for Asian and NHPI populations. These were more than two times greater than other race/ethnicity populations, and more than twenty times greater than estimated for the White population. These patterns were similar for total QALYs lost, which totaled 88,000 (77,000, 100,000) across the study cohort.

**Figure 1.**
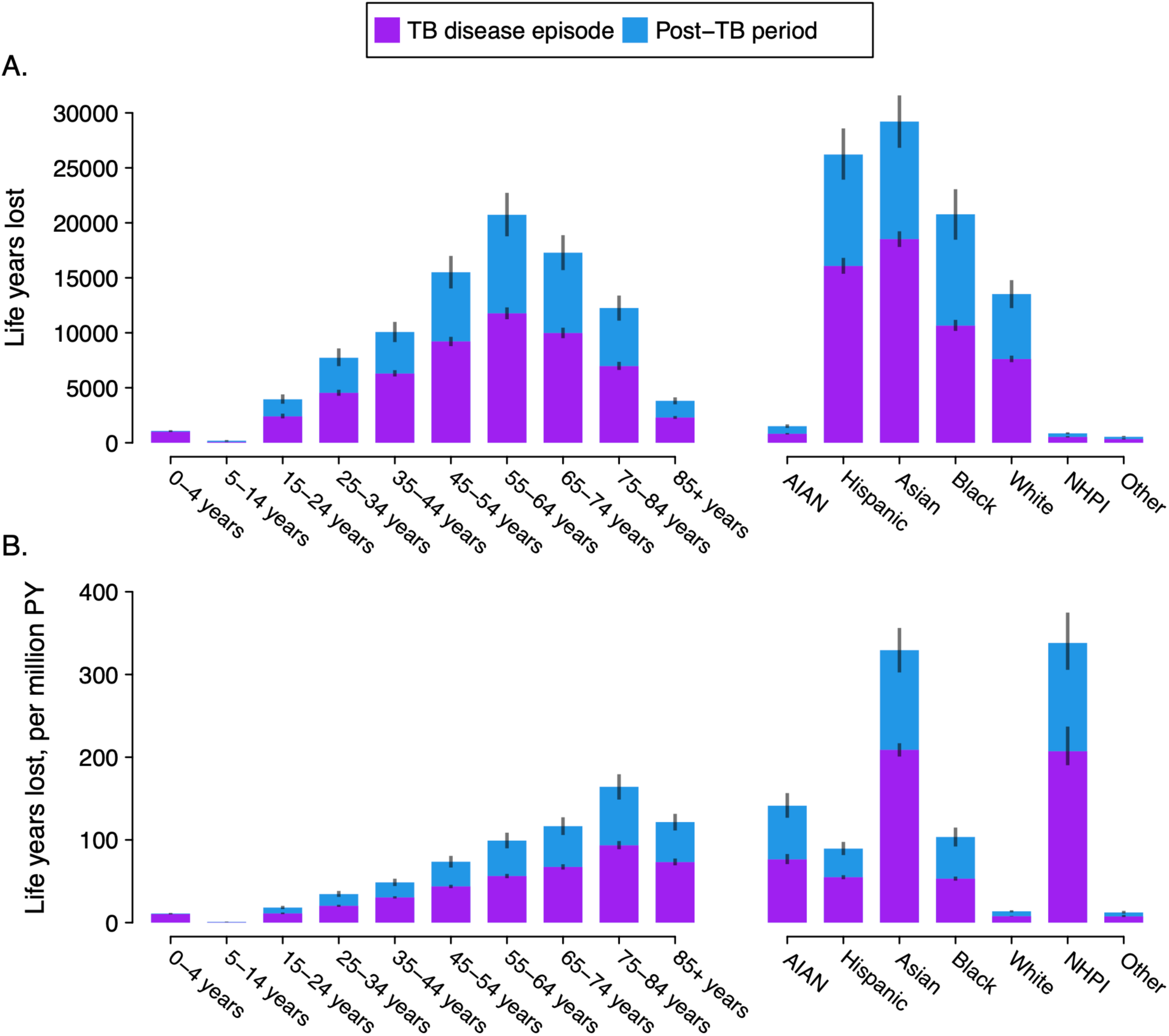
Total life-years lost due to TB in the study cohort, by age group and race/ethnicity. Gray bars represent 95% uncertainty intervals. ‘AIAN’ = American Indian or Alaska Native. ‘Asian’ = Non-Hispanic Asian. ‘Black’ = Non-Hispanic Black. ‘White’ = Non-Hispanic White. ‘NHPI’ = Native Hawaiian or Other Pacific Islander. ‘Other’ = Non-Hispanic other race or people who report more than one race. ‘PY’ = person-years.

### Adjustment for demographic factors

Table 4 reports estimates of how the TB-attributable reduction in life expectancy and quality-adjusted life expectancy varied across each demographic factor, controlling for the others. In these regression analyses, male sex was associated with a 0.16 (0.05, 0.26) greater loss of life-years compared to females, and the 65-74-year-old age group was associated with a greater reduction in life-years compared to other age groups. Across race/ethnicity populations, AIAN race was associated with the greatest reduction in life expectancy, and Asian race the least. Results were similar for regression analyses of QALYs lost.

**Table 4.** Regression estimates of differences in life-years and QALYs lost due to TB for calendar year and demographic factors, by demographic group. QALY = quality-adjusted life-year. ‘AIAN’ = American Indian or Alaska Native. ‘Asian’ = Non-Hispanic Asian. ‘Black’ = Non-Hispanic Black. ‘White’ = Non-Hispanic White. ‘NHPI’ = Native Hawaiian or Other Pacific Islander. ‘Other’ = Non-Hispanic other race or people who report more than one race. Reference group represents category with the greatest number of TB cases in the study sample. Estimates calculated from linear regression model including notification year, sex, age group and race/ethnicity. Regressions also controlled for notification year (non-significant).

|  | Life-years lost |  | QALYs lost |  |
| --- | --- | --- | --- | --- |
|  | Estimate | p-value | Estimate | p-value |
| Intercept | 2.51 (2.21, 2.80) | < 0.001 | 2.15 (1.86, 2.44) | < 0.001 |
| Sex |  |  |  |  |
| Male |  |  |  |  |
| Female | -0.16 (-0.26, -0.05) | 0.004 | -0.11 (-0.19, -0.02) | 0.01 |
| Age group |  |  |  |  |
| 0-4 years | -1.93 (-2.36, -1.49) | < 0.001 | -0.71 (-1.17, -0.24) | 0.003 |
| 5-14 years | -2.69 (-3.14, -2.25) | < 0.001 | -1.47 (-1.92, -1.01) | < 0.001 |
| 15-24 years | -1.94 (-2.22, -1.66) | < 0.001 | -0.97 (-1.28, -0.67) | < 0.001 |
| 25-34 years | -1.78 (-2.02, -1.53) | < 0.001 | -0.96 (-1.21, -0.70) | < 0.001 |
| 35-44 years | -1.25 (-1.48, -1.02) | < 0.001 | -0.68 (-0.89, -0.47) | < 0.001 |
| 45-54 years | -0.51 (-0.70, -0.32) | < 0.001 | -0.25 (-0.40, -0.09) | 0.002 |
| 55-64 years |  |  |  |  |
| 65-74 years | 0.26 (0.07, 0.45) | 0.007 | 0.06 (-0.09, 0.21) | 0.45 |
| 75-84 years | 0.21 (-0.01, 0.42) | 0.07 | -0.08 (-0.27, 0.10) | 0.39 |
| 85+ years | -0.57 (-0.88, -0.26) | < 0.001 | -0.72 (-0.98, -0.47) | < 0.001 |
| Race/ethnicity |  |  |  |  |
| Asian |  |  |  |  |
| AIAN | 1.22 (0.73, 1.71) | < 0.001 | 0.91 (0.51, 1.30) | < 0.001 |
| Black | 0.64 (0.45, 0.82) | < 0.001 | 0.45 (0.30, 0.60) | < 0.001 |
| Hispanic | 0.43 (0.30, 0.57) | < 0.001 | 0.32 (0.21, 0.43) | < 0.001 |
| Other | 0.46 (-0.18, 1.10) | 0.16 | 0.36 (-0.15, 0.86) | 0.17 |
| NHPI | 0.53 (0.05, 1.01) | 0.03 | 0.37 (-0.01, 0.75) | 0.06 |
| White | 0.40 (0.22, 0.58) | < 0.001 | 0.29 (0.15, 0.44) | < 0.001 |

### Sensitivity analyses

The estimated reduction in life expectancy due to TB was 1.74 (1.55, 1.93) years (86% of the main analysis value) when we re-estimated results having omitted individuals with incomplete data (Table S5). When we imputed the best and worst possible outcomes for individuals with missing data on TB survival, the estimated reduction in life expectancy due to TB varied from 1.95 (1.76, 2.13) to 3.75 (3.56, 3.92) years, 97% and 185% of the main analysis value, respectively (Table S6).

## Discussion

In this study, we projected lifetime health outcomes for the 45,738 individuals with reported TB disease in the United States over 2015-2019. These individuals were estimated to lose an average 2.0 life-years, and 1.9 QALYs over their remaining lives as a result of TB disease. These health impacts represent 6.3% and 7.3% reductions in average life expectancy and quality-adjusted life expectancy, respectively. When summed across the study cohort, this represented 93,000 life-years and 88,000 QALYs lost, due to TB. We found a substantial proportion of these health losses—41% for life expectancy, and 48% for quality-adjusted life expectancy—to result from post-TB sequelae. As post-TB health losses have generally been excluded from past analyses of the health impact of TB, the inclusion of post-TB sequelae in the analysis increased the estimated health losses from TB by 70% and 93% for life-years and QALYs respectively (calculated as 1/(1-0.41) and 1/(1-0.48) respectively).

The magnitude of health losses varied across the study cohort. Per person developing TB, the estimated health losses were greatest for 65-74-year-olds. The finding that health losses per TB case peak in older age groups contrasts with the fact that the life-years available to be lost per death decline with age. For example, the life-years lost for a death at age 20 (60 years, based on general population mortality rates)^22^ are 3.7 times greater than the life-years lost for a death at age 70 (16 years). In comparison, life-years lost per person developing TB were estimated to be 3.4 times greater for 65-74-year-olds compared to 15-24-year-olds. The major factor explaining this difference was the case fatality of TB disease, with the proportion of individuals with TB who die before or during treatment rising rapidly with age. In the study cohort (after missing data imputation), 1.1% of 15-24-year-olds died during the TB episode, while for 65-74-year-olds the equivalent value was 16.3%. These results are consistent with prior U.S. studies that have reported rising TB case fatality with age,^26–30^ and evidence of higher TB case fatality at older ages has been available since the early years of TB research.^31^ Despite this, many analyses of TB health consequences have assumed a constant excess mortality rate for TB disease, which would imply that younger individuals with TB experience the greatest health losses. Accurately representing age-specific TB health losses could change the health benefits estimated for TB interventions that deliberately or indirectly target specific age groups (such as household contact tracing, or workplace interventions).

Health losses per TB case were also greater for men vs. women, and for American Indian or Alaska Native persons vs. other race/ethnicity populations. As these populations also experience elevated TB incidence rates,^17^ the differences in health losses per TB case serve to exacerbate TB health disparities for these groups. In contrast, Asian individuals had relatively low number of life-years and QALYs lost per TB case, but were still estimated to represent a major share of overall health losses due to high incidence rates in this population.^17^

Several earlier studies have estimated the lifetime health losses due to TB in U.S. populations, with the average life-years lost per TB case ranging from 1.5 to 7.0 years.^8,27,32,33^ Each of these studies considered specific subnational geographies or trial populations, which may contribute to the variation in reported estimates. In addition, most of these studies used general population life tables to estimate lost life expectancy. While this approach is common, it doesn’t reflect the concentration of mortality risk factors in TB cohorts and will therefore not represent the actual extension in life expectancy had TB not occurred but all other mortality risk factors still been present. All other things being equal, an approach that considers these additional mortality risk factors (as in this analysis) will generate more conservative estimates of life-years lost compared to the approach taken by other analyses.

This study has several limitations. First, we imputed missing values for several covariates used in the analysis. In particular, data on survival of the TB episode was imputed for 6% of individuals. Our imputation approach—assuming survival of these individuals was comparable to survival rates among individuals with similar values for other available covariates—is arguably conservative, as individuals who refuse treatment or are lost-to-follow-up will likely experience worse outcomes than those who follow treatment recommendations. If the difference in survival for these individuals were substantial, our results would underestimate total TB-related health losses. Second, not all deaths in the TB cohort will be attributable to TB, and we were not able to assign each death in the NTSS data to a TB or non-TB cause. Instead, we inferred the additional TB-attributable deaths by comparison to a simulated no-TB counterfactual cohort. In a 2018 study that conducted in-depth record abstraction to classify cause of death in a U.S. TB cohort, 22% of classifiable deaths were attributed to a non-TB cause.^29^ In our study, over the first year of follow-up, estimated mortality risk in the non-TB cohort was 24% of the mortality risk in the TB cohort. This provides some corroboration of the additional TB-associated mortality we estimated for the TB episode. Third, we used mathematical modelling to simulate outcomes during the post-TB period, which introduced uncertainty around parameter values and required several structural assumptions. Notably, we chose to restrict the impact of post-TB on mortality rates to the average follow-up time reported in the source study (6.8 years).^8^ If we had allowed this mortality effect to apply over a longer duration the estimated contribution of post-TB to overall health losses would have been greater. Nationally representative U.S. data on long-term outcomes for TB survivors would be valuable for validating our approach. Fourth, we restricted our study outcomes to life expectancy and QALYs. While evidence on other outcomes (specific health conditions, healthcare utilization, economic consequences) would be valuable, we were not able to identify applicable U.S. data to include these additional outcomes in the analysis. Fifth, the chosen study period (2015-2019) omits the most recent TB data, as treatment outcomes observed during the COVID-19 pandemic may be less generalizable once the healthcare access barriers observed during the peak of the pandemic were resolved.^34^ Consequently, our results might not be applicable to individuals developing TB during 2020-2022 or similar periods of disruption.

Although the United States has been successful at interrupting TB transmission and reducing incidence, TB still represents a major health event for those developing the disease. Even for those who successfully complete treatment, many TB survivors still experience worse health outcomes compared with people who did not have TB. Some of the TB-associated deaths and post-TB sequelae estimated in our analysis are likely avoidable. For example, deaths before diagnosis could be with reduced with broad access to quality healthcare, allowing prompt diagnosis and treatment. Similarly, TB-attributable morbidity and mortality could be reduced if TB treatment were initiated earlier in the course of disease, before substantial organ damage occurs.^35–37^ Alongside interventions that prevent TB occurring in the first place,^38^ efforts to achieve prompt diagnosis and treatment, to support successful treatment completion, and to provide comprehensive medical follow-up for individuals with post-TB sequelae are important for limiting the lifetime health losses caused by TB.

## Funding and disclaimer

This project was funded by the U.S. Centers for Disease Control and Prevention, National Center for HIV, Viral Hepatitis, STD, and TB Prevention Epidemiologic and Economic Modeling Agreement (NEEMA; #1NU38PS004651). The findings and conclusions in this report are those of the authors and do not necessarily represent the views of the Centers for Disease Control and Prevention or other affiliated institutions.

## Data Availability

National TB Surveillance System data contain information abstracted from the national tuberculosis case report form called the Report of Verified Case of Tuberculosis (RVCT) (OMB No. 0920-0728). These data have been reported voluntarily to CDC by state and local health departments and are protected under the Assurance of Confidentiality (Sections 306 and 308(d) of the Public Health Service Act, 42 U.S.C. 242k and 242m(d)), which prevents disclosure of any information that could be used to directly or indirectly identify patients. For more information, see the CDC/ATSDR Policy on Releasing and Sharing Data (at http://www.cdc.gov/maso/Policy/ReleasingData.pdf). A limited dataset is available at http://wonder.cdc.gov/tb.html. Researchers seeking additional National TB Surveillance System data may request access through the National Center for Health Statistics Research Data Centers (https://www.cdc.gov/rdc/b1datatype/tuberculosis.htm).

## Acknowledgements

We thank Carla Winston, Rebekah Stewart, and Joshua Salomon for input on imputation methods and analytic approach. We thank Stephanie Su and Teresa Puente for assistance in study coordination.

## Notes

### Competing Interest Statement

This study was funded by the U.S. Centers for Disease Control and Prevention.

### Author Declarations

The Institutional Review Board (IRB) of the Harvard T.H. Chan School of Public Health determined that this submission is not human subjects research as defined by DHHS regulations or FDA regulations (IRB22-1328).

