## Supplement for "Contribution of post-TB sequelae to life-years and quality-adjusted life-years lost due to TB disease in the United States, 2015-2019"

### Supplementary Methods

Imputation of missing values

Imputation of missing data was implemented using Multivariate Imputation by Chained Equations, via the MICE package in R (v 3.16.0).[1] Values to be imputed were assumed to be missing at random, conditional on the other observed covariates. Using this approach, we created 100 multiply-imputed datasets which were used to represent the uncertainty associated with the imputation.

Uncertainty analysis

We propagated uncertainty through the analysis using 2nd-order Monte Carlo simulation.[2] To do so, we created prior distributions for each uncertain parameter shown in Table 1. Beta distributions were used for parameters bounded [0, 1] (e.g., probabilities, utility weights), and Gamma distributions were used for parameters bounded [0, ∞] (e.g., rates, rate ratios). These distributions were created to have mean values and 95% interval widths matching the values given in Table 1. We drew a sample of 1000 parameter sets using a Latin hypercube sampling approach, and re-estimated the analysis for each of these parameter sets, each time using a randomly-drawn imputed dataset. This procedure produced a sample of 1000 results for each study outcome. We reported point estimates as the mean of these results, and created equal-tailed 95% uncertainty intervals from the 2.5th and 97.5th percentiles of the distribution of results.

Approach for combining utility weights for multiple conditions

We used a multiplicative approach to combine utility weights from multiple conditions:

$$U_{ti} = u_{a+t}^{BG} * u_t^{TB} * \prod_{j \in J} \left(u^j\right)^{I_{ji}}$$

Where $U_{ti}$ represents overall utility for individual $i$ in month $t$, $u_{a+t}^{BG}$ represents background utility as a function of $t$ and starting age $a$, $u_t^{TB}$ is the utility weight from TB in month $t$, $u^j$ is the utility weight for co-prevalent condition $j$, and $I_{ji}$ is an indicator variable equal to 1 when $j$ is present for individual $i$, and 0 otherwise.

Approach for combining mortality risk ratios from multiple co-prevalent conditions

For individuals with multiple co-prevalent conditions we assumed their effect on overall mortality would combine additively:[3,4]

$$mrr_i^C = 1 + \sum_{j \in J} \left( mrr^j - 1 \right) * I_{ji}$$

Where $mrr_i^C$ represents the overall mortality rate ratio for co-prevalent conditions for individual $i$, $mrr^j$ represents the mortality rate ratio for condition $j$, and $I_{ji}$ is an indicator variable equal to 1 when $j$ is present for individual $i$, and otherwise 0.

### Supplementary Exhibits

| | Total cohort | Outcome of TB episode | | |
|---|---|---|---|---|
| | | Died before diagnosis | Died during treatment | Survived |
| Overall | 45,738 (45,738, 45,738) | 1,086 (1,070, 1,105) | 3,201 (3,163, 3,235) | 41,451 (41,409, 41,494) |
| No co-prevalent condition | 27,564 (27,537, 27,587) | 446 (423, 460) | 1,164 (1,147, 1,185) | 25,954 (25,913, 25,992) |
| HIV | 2,346 (2,321, 2,372) | 109 (89, 135) | 232 (218, 245) | 2,005 (1,986, 2,033) |
| Diabetes | 9,054 (9,054, 9,054) | 281 (273, 290) | 1,064 (1,050, 1,081) | 7,708 (7,692, 7,723) |
| ESRD | 1,214 (1,214, 1,214) | 123 (119, 128) | 354 (347, 362) | 737 (727, 746) |
| Organ transplant | 285 (285, 285) | 15 (14, 17) | 47 (45, 51) | 223 (220, 226) |
| Immunosup. meds | 2,727 (2,727, 2,727) | 149 (143, 156) | 489 (478, 503) | 2,089 (2,075, 2,101) |
| MDR-TB | 515 (511, 522) | 7 (6, 10) | 45 (40, 51) | 463 (457, 472) |
| Excess alcohol use | 4,213 (4,192, 4,236) | 151 (140, 163) | 396 (384, 412) | 3,666 (3,643, 3,689) |
| IDU | 595 (587, 606) | 24 (18, 32) | 68 (62, 75) | 502 (490, 514) |
| Experiencing homelessness | 2,183 (2,174, 2,192) | 40 (35, 48) | 204 (193, 218) | 1,939 (1,925, 1,954) |
| Any co-prevalent condition | 18,174 (18,151, 18,201) | 640 (627, 660) | 2,037 (2,008, 2,065) | 15,497 (15,467, 15,536) |

#### Table S1. Study cohort and outcomes of TB disease episode, by presence of co-prevalent conditions.

Results include imputed values for missing demographic variables and treatment outcomes. Values in parentheses represent the range of values across 100 multiply-imputed datasets. 'ESRD' = end-stage renal disease or chronic renal failure. 'Immunosup. meds' = immunosuppressive medications. 'MDR-TB' = multidrug-resistant TB. 'IDU' = injection drug use.

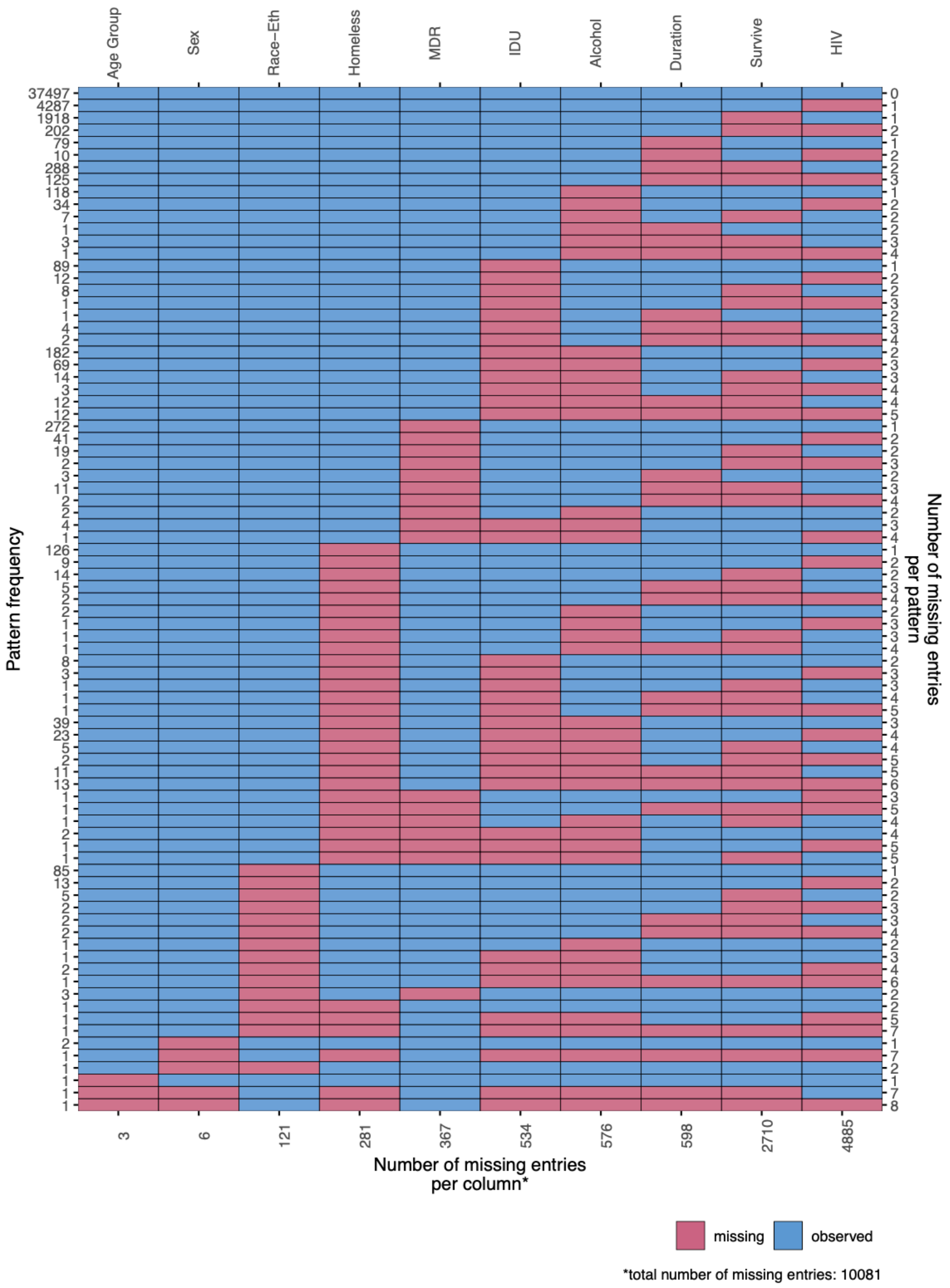

**Figure S1: Missingness pattern for variables derived from the National TB Surveillance System for 2015-2019.***

'Race-Eth' = categorical self-reported race-ethnicity. 'Homeless' (experiencing homelessness at diagnosis) 'MDR' (multidrug-resistant TB), 'IDU' (injection drug use), 'Alcohol' (excess alcohol use), and 'HIV' = binary variables for presence/absence of co-prevalent conditions. 'Duration' = treatment duration in months. 'Survive' = categorical variable for survival of TB episode (categories 'dead at diagnosis', 'died on treatment', and 'survived'). Variables describing notification year, organ transplant receipt, ESRD (end-stage renal disease or chronic renal failure), diabetes, and receipt of immunosuppressive medications had no missing values and are excluded from figure.

| | TB cohort | No TB counterfactual | Percentage reduction due to TB |
|---|---|---|---|
| Overall | 30.3 (29.9, 30.7) | 32.3 (31.9, 32.7) | 6.3 (5.7, 6.8) |
| Sex | | | |
| Men | 27.8 (27.4, 28.3) | 30.0 (29.6, 30.4) | 7.2 (6.5, 7.8) |
| Women | 34.0 (33.7, 34.3) | 35.8 (35.5, 36.1) | 5.1 (4.7, 5.6) |
| Age group | | | |
| 0-4 years | 75.7 (75.6, 75.8) | 76.7 (76.6, 76.8) | 1.3 (1.2, 1.4) |
| 5-14 years | 68.3 (68.2, 68.4) | 68.5 (68.4, 68.6) | 0.3 (0.3, 0.4) |
| 15-24 years | 57.5 (57.4, 57.7) | 58.4 (58.3, 58.6) | 1.5 (1.4, 1.7) |
| 25-34 years | 48.1 (47.9, 48.4) | 49.2 (48.9, 49.4) | 2.1 (1.9, 2.3) |
| 35-44 years | 36.8 (36.4, 37.3) | 38.4 (38.0, 38.8) | 4.1 (3.8, 4.5) |
| 45-54 years | 25.8 (25.2, 26.4) | 28.1 (27.6, 28.7) | 8.3 (7.5, 9.1) |
| 55-64 years | 17.7 (17.1, 18.3) | 20.5 (19.9, 21.1) | 13.8 (12.5, 15.2) |
| 65-74 years | 11.4 (10.8, 11.9) | 14.4 (13.9, 14.9) | 21.1 (19.2, 23.1) |
| 75-84 years | 5.9 (5.5, 6.2) | 8.8 (8.5, 9.1) | 33.5 (30.6, 36.5) |
| 85+ years | 2.5 (2.3, 2.6) | 4.6 (4.5, 4.8) | 47.1 (43.8, 50.3) |
| Race/ethnicity | | | |
| AIAN | 25.3 (24.9, 25.9) | 28.2 (27.7, 28.6) | 10.1 (9.0, 11.1) |
| Hispanic | 33.9 (33.4, 34.4) | 35.9 (35.5, 36.3) | 5.5 (5.1, 6.0) |
| Asian | 31.6 (31.3, 32.0) | 33.4 (33.1, 33.8) | 5.4 (5.0, 5.8) |
| Black | 28.0 (27.6, 28.4) | 30.2 (29.8, 30.5) | 7.3 (6.5, 8.1) |
| White | 20.8 (20.4, 21.1) | 23.2 (22.9, 23.5) | 10.5 (9.5, 11.5) |
| NHPI | 41.5 (41.0, 42.1) | 43.2 (42.8, 43.8) | 3.9 (3.6, 4.3) |
| Other | 35.5 (35.1, 35.8) | 37.3 (37.0, 37.7) | 5.1 (4.5, 5.9) |

### Table S2. Estimated life expectancy by scenario, notification year and demographic group.

Values in parentheses represent 95% uncertainty intervals. QALE = quality-adjusted life expectancy. ‘AIAN’ = American Indian or Alaska Native. ‘Asian’ = Non-Hispanic Asian. ‘Black’ = Non-Hispanic Black. ‘White’ = Non-Hispanic White. ‘NHPI’ = Native Hawaiian or Other Pacific Islander. ‘Other’ = Non-Hispanic other race or people who report more than one race.

| | TB cohort | No TB counterfactual | Percentage reduction due to TB |
|---|---|---|---|
| Overall | 24.5 (23.1, 25.7) | 26.4 (24.9, 27.6) | 7.3 (6.5, 8.1) |
| Sex | | | |
| Men | 22.5 (21.2, 23.6) | 24.5 (23.2, 25.7) | 8.2 (7.4, 9.1) |
| Women | 27.5 (25.9, 28.8) | 29.3 (27.7, 30.7) | 6.2 (5.5, 6.9) |
| Age group | | | |
| 0-4 years | 65.3 (62.8, 67.5) | 67.1 (64.6, 69.3) | 2.6 (2.1, 3.3) |
| 5-14 years | 58.1 (55.7, 60.2) | 59.1 (56.7, 61.2) | 1.7 (1.1, 2.3) |
| 15-24 years | 48.1 (45.8, 50.1) | 49.5 (47.2, 51.6) | 2.9 (2.3, 3.6) |
| 25-34 years | 39.5 (37.5, 41.3) | 40.9 (38.9, 42.8) | 3.5 (2.9, 4.2) |
| 35-44 years | 29.6 (27.9, 31.1) | 31.3 (29.6, 32.8) | 5.5 (4.8, 6.2) |
| 45-54 years | 20.2 (18.9, 21.4) | 22.3 (20.9, 23.6) | 9.6 (8.7, 10.7) |
| 55-64 years | 13.5 (12.5, 14.4) | 15.9 (14.7, 16.9) | 15.0 (13.6, 16.6) |
| 65-74 years | 8.4 (7.7, 9.1) | 10.8 (9.9, 11.7) | 22.3 (20.3, 24.3) |
| 75-84 years | 4.2 (3.8, 4.6) | 6.4 (5.8, 7.0) | 34.9 (31.9, 37.8) |
| 85+ years | 1.7 (1.5, 1.9) | 3.3 (2.9, 3.6) | 49.1 (45.8, 52.3) |
| Race/ethnicity | | | |
| AIAN | 21.0 (19.9, 22.0) | 23.6 (22.3, 24.6) | 10.9 (9.8, 12.1) |
| Hispanic | 27.5 (26.1, 28.9) | 29.5 (27.9, 30.9) | 6.6 (5.9, 7.4) |
| Asian | 25.2 (23.7, 26.6) | 27.0 (25.4, 28.3) | 6.4 (5.7, 7.2) |
| Black | 22.9 (21.8, 24.0) | 25.0 (23.8, 26.2) | 8.3 (7.4, 9.3) |
| White | 16.6 (15.6, 17.5) | 18.7 (17.6, 19.7) | 11.4 (10.3, 12.5) |
| NHPI | 34.5 (32.8, 36.0) | 36.3 (34.6, 37.9) | 5.0 (4.4, 5.8) |
| Other | 29.1 (27.5, 30.5) | 31.1 (29.5, 32.4) | 6.2 (5.4, 7.2) |

**Table S3. Estimated quality-adjusted life expectancy by scenario, notification year and demographic group.**

Values in parentheses represent 95% uncertainty intervals. QALE = quality-adjusted life expectancy. ‘AIAN’ = American Indian or Alaska Native. ‘Asian’ = Non-Hispanic Asian. ‘Black’ = Non-Hispanic Black. ‘White’ = Non-Hispanic White. ‘NHPI’ = Native Hawaiian or Other Pacific Islander. ‘Other’ = Non-Hispanic other race or people who report more than one race.

| | Life-years lost | | QALYs lost | |
|---|---|---|---|---|
| | Total | Per million person-years | Total | Per million person-years |
| Overall | 92,639 (84,364, 101,079) | 57 (52, 62) | 88,146 (77,238, 99,730) | 54 (48, 61) |
| Sex | | | | |
| Men | 59,731 (54,291, 65,290) | 75 (68, 82) | 55,611 (48,797, 62,714) | 69 (61, 78) |
| Women | 32,907 (30,061, 35,776) | 40 (36, 43) | 32,535 (28,341, 36,978) | 39 (34, 45) |
| Age group | | | | |
| 0-4 years | 1,082 (1,040, 1,152) | 11 (11, 12) | 1,911 (1,521, 2,415) | 19 (16, 25) |
| 5-14 years | 190 (159, 250) | 1 (1, 1) | 879 (597, 1,241) | 4 (3, 6) |
| 15-24 years | 3,953 (3,548, 4,397) | 18 (16, 20) | 6,284 (4,987, 7,778) | 29 (23, 36) |
| 25-34 years | 7,732 (6,967, 8,583) | 35 (31, 38) | 10,575 (8,722, 12,759) | 47 (39, 57) |
| 35-44 years | 10,077 (9,156, 10,991) | 49 (44, 53) | 10,890 (9,470, 12,470) | 53 (46, 60) |
| 45-54 years | 15,509 (14,033, 16,991) | 74 (67, 81) | 14,260 (12,567, 16,005) | 68 (60, 76) |
| 55-64 years | 20,735 (18,775, 22,726) | 99 (90, 109) | 17,491 (15,452, 19,649) | 84 (74, 94) |
| 65-74 years | 17,288 (15,698, 18,877) | 117 (106, 127) | 13,708 (12,004, 15,343) | 92 (81, 104) |
| 75-84 years | 12,257 (11,103, 13,388) | 164 (149, 179) | 9,329 (8,134, 10,464) | 125 (109, 140) |
| 85+ years | 3,816 (3,495, 4,127) | 122 (111, 132) | 2,818 (2,455, 3,178) | 90 (78, 101) |
| Race/ethnicity | | | | |
| AIAN | 1,510 (1,354, 1,672) | 141 (127, 157) | 1,368 (1,205, 1,529) | 128 (113, 143) |
| Hispanic | 26,217 (23,925, 28,587) | 89 (82, 98) | 25,761 (22,564, 29,105) | 88 (77, 99) |
| Asian | 29,211 (26,821, 31,590) | 329 (302, 356) | 27,994 (24,446, 31,829) | 316 (276, 359) |
| Black | 20,778 (18,464, 23,058) | 104 (92, 115) | 19,710 (17,222, 22,344) | 98 (86, 111) |
| White | 13,524 (12,245, 14,789) | 14 (12, 15) | 11,831 (10,406, 13,268) | 12 (11, 13) |
| NHPI | 858 (776, 951) | 338 (306, 375) | 928 (800, 1,074) | 366 (315, 423) |
| Other | 541 (484, 628) | 12 (11, 14) | 554 (477, 643) | 13 (11, 15) |

#### Table S4 Total life-years and quality-adjusted life-years lost due to TB in the study cohort, by notification year and demographic group.

Values in parentheses represent 95% uncertainty intervals. QALY = quality-adjusted life-year. ‘AIAN’ = American Indian or Alaska Native. ‘Asian’ = Non-Hispanic Asian. ‘Black’ = Non-Hispanic Black. ‘White’ = Non-Hispanic White. ‘NHPI’ = Native Hawaiian or Other Pacific Islander. ‘Other’ = Non-Hispanic other race or people who report more than one race.

| | Reduction in life expectancy | | Reduction in quality-adjusted life expectancy | |
|---|---|---|---|---|
| | Main analysis | Complete case analysis | Main analysis | Complete case analysis |
| Overall | 2.03 (1.84, 2.21) | 1.74 (1.55, 1.93) | 1.93 (1.69, 2.18) | 1.72 (1.49, 1.97) |
| Sex | | | | |
| Men | 2.15 (1.96, 2.35) | 1.88 (1.67, 2.09) | 2.00 (1.76, 2.26) | 1.80 (1.57, 2.06) |
| Women | 1.83 (1.67, 1.99) | 1.53 (1.37, 1.69) | 1.81 (1.58, 2.06) | 1.59 (1.36, 1.85) |
| Age group | | | | |
| 0-4 years | 0.99 (0.95, 1.05) | 0.56 (0.54, 0.57) | 1.75 (1.39, 2.21) | 1.37 (1.02, 1.83) |
| 5-14 years | 0.21 (0.18, 0.28) | 0.14 (0.10, 0.17) | 0.98 (0.66, 1.38) | 0.91 (0.60, 1.31) |
| 15-24 years | 0.89 (0.80, 0.99) | 0.75 (0.67, 0.83) | 1.42 (1.13, 1.76) | 1.30 (1.02, 1.64) |
| 25-34 years | 1.04 (0.94, 1.15) | 0.91 (0.81, 1.01) | 1.42 (1.17, 1.71) | 1.32 (1.07, 1.60) |
| 35-44 years | 1.59 (1.45, 1.74) | 1.41 (1.27, 1.55) | 1.72 (1.50, 1.97) | 1.57 (1.34, 1.82) |
| 45-54 years | 2.34 (2.12, 2.57) | 2.08 (1.86, 2.30) | 2.15 (1.90, 2.42) | 1.95 (1.72, 2.21) |
| 55-64 years | 2.83 (2.56, 3.10) | 2.45 (2.17, 2.73) | 2.38 (2.11, 2.68) | 2.11 (1.84, 2.40) |
| 65-74 years | 3.04 (2.76, 3.32) | 2.61 (2.32, 2.90) | 2.41 (2.11, 2.70) | 2.10 (1.82, 2.37) |
| 75-84 years | 2.95 (2.67, 3.23) | 2.54 (2.25, 2.83) | 2.25 (1.96, 2.52) | 1.96 (1.67, 2.23) |
| 85+ years | 2.19 (2.00, 2.36) | 1.84 (1.64, 2.03) | 1.61 (1.41, 1.82) | 1.38 (1.18, 1.58) |
| Race/ethnicity | | | | |
| AIAN | 2.83 (2.54, 3.13) | 2.45 (2.14, 2.75) | 2.56 (2.26, 2.87) | 2.24 (1.95, 2.56) |
| Hispanic | 1.99 (1.81, 2.17) | 1.69 (1.52, 1.87) | 1.95 (1.71, 2.21) | 1.73 (1.49, 1.99) |
| Asian | 1.80 (1.66, 1.95) | 1.48 (1.33, 1.62) | 1.73 (1.51, 1.97) | 1.50 (1.29, 1.73) |
| Black | 2.19 (1.95, 2.44) | 2.01 (1.77, 2.26) | 2.08 (1.82, 2.36) | 1.95 (1.69, 2.23) |
| White | 2.43 (2.20, 2.66) | 2.10 (1.86, 2.35) | 2.13 (1.87, 2.38) | 1.89 (1.64, 2.15) |
| NHPI | 1.70 (1.54, 1.88) | 1.51 (1.36, 1.66) | 1.83 (1.58, 2.12) | 1.68 (1.44, 1.95) |
| Other | 1.89 (1.69, 2.19) | 1.45 (1.29, 1.62) | 1.93 (1.67, 2.24) | 1.56 (1.32, 1.82) |

**Table S5. Average reduction in life expectancy and quality-adjusted life expectancy for individuals developing TB, main analysis versus complete case analysis.**

Values in parentheses represent 95% uncertainty intervals. QALE = quality-adjusted life expectancy. ‘AIAN’ = American Indian or Alaska Native. ‘Asian’ = Non-Hispanic Asian. ‘Black’ = Non-Hispanic Black. ‘White’ = Non-Hispanic White. ‘NHPI’ = Native Hawaiian or Other Pacific Islander. ‘Other’ = Non-Hispanic other race or people who report more than one race.

| | Reduction in life expectancy | | Reduction in quality-adjusted life expectancy | |
|---|---|---|---|---|
| | Optimistic imputation | Pessimistic imputation | Optimistic imputation | Pessimistic imputation |
| Overall | 1.95 (1.76, 2.13) | 3.75 (3.56, 3.92) | 1.87 (1.63, 2.12) | 3.32 (3.04, 3.61) |
| Sex | | | | |
| Men | 2.07 (1.87, 2.27) | 4.05 (3.86, 4.24) | 1.94 (1.70, 2.19) | 3.55 (3.25, 3.84) |
| Women | 1.77 (1.61, 1.92) | 3.27 (3.11, 3.42) | 1.76 (1.53, 2.01) | 2.97 (2.70, 3.24) |
| Age group | | | | |
| 0-4 years | 0.97 (0.95, 0.99) | 2.13 (2.11, 2.18) | 1.73 (1.38, 2.19) | 2.74 (2.38, 3.18) |
| 5-14 years | 0.21 (0.18, 0.24) | 1.49 (1.46, 1.54) | 0.97 (0.66, 1.38) | 2.06 (1.74, 2.45) |
| 15-24 years | 0.85 (0.76, 0.93) | 4.84 (4.77, 4.92) | 1.38 (1.10, 1.71) | 4.73 (4.39, 5.08) |
| 25-34 years | 0.98 (0.88, 1.09) | 4.34 (4.24, 4.43) | 1.38 (1.13, 1.66) | 4.13 (3.84, 4.44) |
| 35-44 years | 1.52 (1.38, 1.66) | 3.63 (3.49, 3.78) | 1.66 (1.43, 1.91) | 3.37 (3.10, 3.65) |
| 45-54 years | 2.25 (2.03, 2.48) | 3.71 (3.50, 3.92) | 2.08 (1.84, 2.34) | 3.23 (2.95, 3.52) |
| 55-64 years | 2.72 (2.45, 3.00) | 3.78 (3.52, 4.03) | 2.31 (2.03, 2.60) | 3.12 (2.80, 3.42) |
| 65-74 years | 2.93 (2.65, 3.21) | 3.73 (3.45, 4.01) | 2.33 (2.04, 2.61) | 2.92 (2.61, 3.22) |
| 75-84 years | 2.87 (2.59, 3.15) | 3.23 (2.96, 3.50) | 2.19 (1.91, 2.46) | 2.45 (2.16, 2.74) |
| 85+ years | 2.13 (1.95, 2.31) | 2.28 (2.10, 2.45) | 1.57 (1.36, 1.78) | 1.68 (1.47, 1.89) |
| Race/ethnicity | | | | |
| AIAN | 2.77 (2.48, 3.06) | 3.79 (3.51, 4.08) | 2.52 (2.21, 2.83) | 3.36 (3.02, 3.68) |
| Hispanic | 1.89 (1.72, 2.07) | 4.54 (4.37, 4.71) | 1.88 (1.64, 2.14) | 4.04 (3.73, 4.35) |
| Asian | 1.74 (1.59, 1.88) | 3.28 (3.14, 3.43) | 1.68 (1.46, 1.91) | 2.91 (2.65, 3.17) |
| Black | 2.12 (1.88, 2.36) | 3.58 (3.34, 3.80) | 2.02 (1.77, 2.30) | 3.21 (2.92, 3.50) |
| White | 2.35 (2.12, 2.58) | 3.50 (3.29, 3.72) | 2.07 (1.81, 2.33) | 2.99 (2.70, 3.27) |
| NHPI | 1.66 (1.51, 1.82) | 3.24 (3.09, 3.42) | 1.81 (1.56, 2.08) | 3.11 (2.83, 3.42) |
| Other | 1.82 (1.64, 2.00) | 4.25 (4.06, 4.47) | 1.88 (1.64, 2.15) | 3.88 (3.57, 4.19) |

**Table S6 Average reduction in life expectancy and quality-adjusted life expectancy for individuals developing TB, main analysis versus optimistic and pessimistic outcome imputation.**

Values in parentheses represent 95% uncertainty intervals. QALE = quality-adjusted life expectancy. 'AIAN' = American Indian or Alaska Native. 'Asian' = Non-Hispanic Asian. 'Black' = Non-Hispanic Black. 'White' = Non-Hispanic White. 'NHPI' = Native Hawaiian or Other Pacific Islander. 'Other' = Non-Hispanic other race or people who report more than one race.
